# Further Evidence of a Possible Correlation Between the Severity of Covid-19 and BCG Immunization

**DOI:** 10.1101/2020.04.07.20056994

**Authors:** Serge Dolgikh

**Affiliations:** National Aviation University

**Keywords:** Infectious epidemiology, Covid-19

## Abstract

In this work we observe a number of cases supporting the possible correlation between the administration of BCG tuberculosis vaccine and the severity of Covid-19 effects in the population proposed in the earlier works [1]. Based on the early preliminary analysis of the publicly available data we propose a number of arguments and observations providing further support for the correlation hypothesis and make an observation that the effectiveness of the protection effect of BCG immunization, if confirmed, may depend to a significant extent on the age of administration, with the early age inoculation more effective for the lasting protection.

## 1 Introduction

### 1.1 Purpose

A significant variation in the patterns of onset of Covid-19 epidemics across jurisdictions has been noticed previously. Quite pronounced is the difference between the “avalanche” scenario seen in Italy, Spain, France and USA on one hand, and the “mild” one (Taiwan, Germany, many East European jurisdictions and others) on the other. While multiple factors certainly play role in these variations, some can be more influential on the overall dynamics of the impacts of the epidemics.

A possible link between the effects of Covid-19 pandemics such as the rate of spread and the severity of cases; and a universal immunization program against tuberculosis with BCG vaccine (UIP, hereinafter) was suggested in [1]. In this work we provide several additional arguments supporting the case for such correlation based on the available data in the public domain. Further analysis of statistical data is needed to achieve a confident conclusion.

Based on the preliminary analysis of the data we make an important observation that the effectiveness of the protection provided by BCG vaccination against Covid-19 may extend more or substantially more to the cases where it is provided at birth or young age.

## 2 Data

### 2.1 Terminology

Time aspect is critically important in this analysis. We propose to define the beginning moment of the developments related to Covid-19 globally as the date of the first official release of information about Covid-19 on the 31.12.2019 [2], though the first cases of the epidemics including the zero event of possible animal to human crossover almost certainly happened earlier.

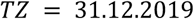

Along with the global Time Zero we also suggest to define the local Time Zero (LTZ) indicating the time of arrival of the epidemics in the given locality. It can be sensibly defined as the date of the first confirmed case in the area.

Two common measures of the epidemics impact on the human population used throughout: Cases per capita (c.p.c.) and Mortality per capita (m.p.c):

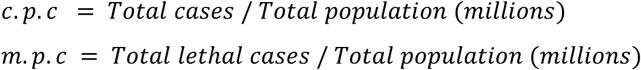

Note that the former measure strongly depends on the methods and policies in the reporting jurisdiction such as testing strategy, verification of reported cases and so on. Also, it’s not always possible to verify the difference between suspected or reported cases, and those that were later confirmed as Covid-19. For that reason, we suggest it to be used as a trend indicative parameter and more often used m.p.c. as the current and more accurate measure of the epidemics impact, on the assumption that policies and protocols in the selected administration allow more accurate identification of cause and reporting.

### 2.2 Case Data

Here we attempted to collect data from a number of jurisdictions as an input to the analysis of the significance of the correlation hypothesis. A comprehensive statistical study to this effect should be undertaken, for which the presented data can provide a convincing rationale, but hardly a replacement.

Vaccination level is measured in the following bands as defined in [3]:

A: has universal or near-universal BCG coverage

B: had BCG vaccination in the past covering significant part of population (> 50%)

B2: never had a universal program, but some coverage (e.g. regional, specific groups)

C: never had a UIP/BCG

We define three groups of countries, by the reported impact of Covid-19 on the population:

Very Low (VL), m.p.c. < 1 or near

Low (L): 2 < m.p.c. < 10 or near

High (H): m.p.c. near or greater than 100

The data <u>was not</u> selected to fit the hypothesis. Rather, several common sense criteria were applied such as: certain expectation of reliability and consistency of the reporting jurisdiction; a reasonable level of exposure to Covid-19 e.g. certain minimum number of reported cases; geographical and development level variation; and the well-known cases that already received attention. A comprehensive and more formal statistical analysis will be the subject of a future study.

### Cautions

- Consistency and reliability of data reported by the national, regional and local health administrations.
- Alignment in time of beginning of epidemics: the situation is developing quickly and the phases of epidemics development may not be aligned.
- Alignment in the time of reporting may be an issue due to reporting practices of jurisdictions.
- Especially the availability, consistency and reliability of historical data and statistics on the administration of immunization programs in the national, regional and so on, jurisdictions can be an issue.

## 3 Early Observations

### 3.1 UIP Correlation Hypothesis

While fully aware about the concerns and caveats regarding the integrity of data used in these notes, we believe that the developing situation does not leave the option for a prolonged wait for availability of better data as grounded decisions need to be taken daily. While the work on collecting the data has to continue, some initial conclusions need to be derived in an ongoing manner, while constantly checked and verified against more confident data.

#### Known factors

Time of the introduction

Demographics

Social, tradition, lifestyle

Economic and social development

Quality and efficiency of the healthcare system

Quality of policy making and execution

#### Hypothesis

The hypothesis that was proposed in [1] and is investigated here is the presence of a population-wide, or wide-spread factor (F) that influences the dynamics and the outcomes of the epidemics to a significant and measurable extent affecting both rate of spread and severity. This factor is different from the already known factors, listed above.

The examples of the extra factor can be: genetic differences; past widespread public policy such as immunization (a side effect); other yet unknown factors; a combination of multiple factors.

#### Assumptions

We will assume that the behavior and properties of the contagion agent do not vary significantly regionally and globally, and remain relatively stable over the period of the analysis.

### 3.2 Observations

#### 1. Effective management of Covid-19 is possible

A group of countries can be identified where even within the countries of the first wave (see below) the spread of the epidemics has been stabilized and remains on the low level (countries of the group VL: m.p.c. rate below or near 1% of the reported cases).

Not an isolated case, so an analysis of influencing factors is possible.

Includes several countries of the first wave (below), so not explained by a delayed exposure.

Includes a large proportion of countries with large aged demographics, so cannot be explained by the demographics only.

Includes several countries in lower group of economic development however, these cases may still be in the developing phase.

#### 2. Multiple waves

Two broad group of cases can be identified from the data in Table 1:

**Table 1.** Covid-19 Impacts by Jurisdiction (updated 07.04.2020)

| *Case* | *Popula-*<br>*tion (M)* | *Start <sup>(1)</sup>* | *Cases,*<br>*tot* | *Mort.,*<br>*tot* | *Cases,*<br>*cap* | *Mort.,*<br>*cap* | *BCG*<br>*level* |
| --- | --- | --- | --- | --- | --- | --- | --- |
| **VLow** |  |  |  |  |  |  |  |
| Slovakia | 5.5 | 06.03 | 534 | 2 | 85.2 | 0.37 | A |
| Taiwan | 23.8 | 21.01 | 373 | 5 | 15.04 | 0.21 | A |
| Japan | 126.8 | 16.01 | 3664 | 73 | 23.3 | 0.58 | A |
| Ukraine | 42.2 | 29.02 | 1467 | 46 | 29.0 | 1.10 | A |
| Kyiv | 3.8 | 16.03 | 267 | 4 | 64.5 | 1.05 | A |
| Singapore | 5.6 | 23.01 | 1375 | 6 | 208.5 | 1.10 | A |
| **Low** |  |  |  |  |  |  |  |
| Chile | 18.1 | 03.03 | 4815 | 37 | 252 | 2.0 | A |
| Australia | 24.6 | 25.01 | 5899 | 48 | 216.2 | 1.95 | B |
| South Ko-<br>rea | 51.5 | 20.01 | 10331 | 192 | 196.1 | 3.7 | A |
| Moldova | 3.6 | 07.03 | 965 | 19 | 280.4 | 5.3 | A |
| Ukraine,<br>west <sup>(2)</sup> | 2.1 | 03.03 | 380 | 8 | 162.4 | 3.8 | A |
| Israel | 8.7 | 21.02 | 8904 | 57 | 826.7 | 6.6 | B |
| Poland | 38.0 | 04.03 | 4413 | 107 |  | 2.8 | A |
| Czechia | 10.7 | 01.03 | 4362 | 78 | 407.9 | 7.3 | A |
| Croatia | 4.1 | 25.02 | 1222 | 16 | 299 | 3.9 |  |
| Albania | 2.9 | 08.03 | 377 | 21 | 117 | 7.2 | A |
| Greece | 10.5 | 28.02 | 1755 | 79 | 167.1 | 7.5 | A |
| Jordan | 10.1 | 02.03 | 349 | 6 | 32.8 | 0.59 |  |
| Finland | 5.5 | 29.01 | 1875 | 47 | 407.6 | 4.6 | B |
| Canada | 37.6 | 25.01 | 16663 | 323 | 369.8 | 8.6 | B2 |
| Prairies<br>(Can) <sup>(3)</sup> | 2.6 | 12.03 | 425 | 5 | 163.5 | 1.92 | B2<sup>(4)</sup> |
| Ontario (Can) | 14.6 | 25.01 | 4347 | 132 | 248.6 | 9.0 | B2<sup>(4)</sup> |
| Quebec (Can) | 8.5 | 28.02 | 8580 | 121 | 823.2 | 14.2 | B2<sup>(4,5)</sup> |
| Argentina | 45.2 | 03.03 | 1554 | 46 |  | 1.0 | A |
| **Mid** |  |  |  |  |  |  |  |
| Norway | 5.4 | 26.02 | 5551 | 77 | 1028.0 | 14.3 | B |
| Denmark | 5.6 | 27.02 | 4681 | 187 | 85.2 | 33.4 | B |
| Sweden | 10.1 | 31.01 | 6558 | 506 | 654.6 | 50.1 | B |
| Austria | 8.8 | 25.02 | 12297 | 220 |  | 25 | B |
| Germany | 82.8 | 27.01 | 103375 | 1810 | 1113 | 21.9 | B |
| Portugal | 10.3 | 02.03 | 11730 | 311 | 1024 | 30.2 | A |
| Ireland | 4.9 | 29.02 | 5364 | 174 |  | 35.5 | A |
| **High** |  |  |  |  |  |  |  |
| Italy | 60.5 | 31.01 | 124632 | 16523 | 2069 | 273.1 | C |
| Spain <sup>(5)</sup> | 46.7 | 31.01 | 124736 | 11814 | 2648 | 253.0 | B2 |
| UK <sup>(6)</sup> | 66.4 | 29.01 | 51608 | 5373 | 630.7 | 80.9 | B |
| Netherlands | 17.2 | 27.02 | 18803 | 1867 | 952.8 | 108.5 | C |
| Belgium | 11.4 |  | 18431 | 1632 | 1600 | 143.1 | C |
| France <sup>(6,7)</sup> | 67.0 | 24.01 | 74390 | 8911 | 959.2 | 133.0 | B |
| New York | 8.6 | 01.03 | 60850 | 2254 | 7075.6 | 262.1 | C |
| **Other** |  |  |  |  |  |  |  |
| USA | 327.2 | 21.01 | 367461 | 10910 | 909.9 | 33.3 | C |

<sup>1</sup> The first confirmed case of Covid-19, LTZ
<sup>(2)</sup> Ternopil'ska, Chernovits'ka regions (oblast)
<sup>(3)</sup> Manitoba, Saskatchewan provinces
<sup>(4)</sup> Used in targeted communities with higher incidence
<sup>(5)</sup> Limited age cohort (Quebec: 19 years, Spain: 16 years)
<sup>(6)</sup> Immunization started at later age (not at birth)
<sup>(7)</sup> Contradictory data on immunization practice (at birth or later age).

<u>The first wave</u> (∼TZ + 1m): in the month of January, 2020 usually with: direct links to Wuhan in China.

<u>The second wave</u> (∼TZ + 2m): second half of February – early March, prevailing travels to the countries affected by the first wave.

There are reports nowadays of the reflection or tertiary waves reaching the countries of the first wave that have already passed the peak of the epidemics.

#### 3. “Avalanche” and “Mild” scenarios

The scenario with a rapid onset of large number of cases and high mortality (“Avalanche”) has been observed in several jurisdictions of the first wave (above) and in our view, can now be confirmed.

Given a number of instances in both waves we believe that one can confirm the second, managed or “mild” scenario as well, with much slower growth in both cases and mortality. In several jurisdictions of the first wave the latter appears to have been stabilized on the level far below that exhibited by the cases of the first scenario.

Several jurisdictions in this group (VL and L groups in the table) can still be in the developing phase as of the time of writing (TZ+3m). These can be most interesting to observe with respect to the factor F hypothesis when they reach the onset timescale of LTZ + 1-1.5m (i.e. approx. TZ+ 3.5m – 4m), as several of these countries fall into the significantly lower category by economic development (Ukraine, Moldova, Albania) and the current level of epidemics impact, if maintained, would not be possible to explain only by the level of development, the effectiveness of the healthcare system and/or epidemics management policy.

#### 4. The period of the onset

The onset of a mass spread in the population (in the “avalanche” scenario) appears to be 4 to 6 weeks from the first exposure, without strong suppressing measures.

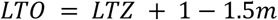

Confirmed by the following cases: Italy, Spain, France, USA, UK (first wave); Netherlands, Norway, Ukraine (2^nd^ wave) and others.

The development of the epidemics in the initial phase may depend on many factors, leading to the relation between the actual introduction of the contagion and the first reported case not necessarily being clear and strong. For example, several cases can go unnoticed until a case with more serious course happens and is brought to the attention of health authorities (and even then, not misdiagnosed). It could mean that even in the countries with the same starting time, the patterns of the development can still be shifted by certain time delta.

#### 5. Clear evidence of a correlation between the severity of Covid-19 and universal immunization program with BCG

All countries in the VL category currently have a UIP or had it recently still covering most of age cohorts.

Majority of countries in the L category have an ongoing UIP or had it relatively recently or in limited use (e.g. Canada, Norway for targeted groups).

No countries from group C (never had a UIP or significant BCG immunization program) are found in the groups VL, L with lower impacts of Covid-19 though regional variations are possible (e.g. Canada).

In some cases of the group H that had a vaccination program at some point, the lower effectiveness can be explained by a conjecture that the protection against Covid-19 has higher effectiveness if the vaccine is given at birth or at a young age (see the next observation).

#### 6. Early age effectiveness hypothesis

As discussed further in Section 4.2 the early results are consistent with the hypothesis that a UIP/BCG is offering some level of protection against the spread and severity of Covid-19 and that the effectiveness of this protection may depend on the age of administration of the vaccine, with the early age policy more effective for a long-term protection.

Table 2 lists the subset of the cases in the immunization band B, where universal immunization was provided but ceased in the past. Note very close correlation between the time since termination of UIP and the m.p.c value of Covid-19 impact.

**Table 2.** UIP termination vs. Covid-19 M.p.c. correlation

| Case | Population | LTZ | M.p.c. | BCG Band | Cessation of UIP |
| --- | --- | --- | --- | --- | --- |
| Sweden | 10.1 | 31.01 | 58.51 | B | 1975 |
| Australia | 24.6 | 25.01 | 1.95 | B | 1985 |
| Denmark * | 5.6 | 27.02 | 36.25 | B | 1986 |
| Israel * | 8.7 | 21.02 | 6.90 | B | 1986 |
| Austria * | 8.8 | 25.02 | 27.61 | B | 1990 |
| Norway * | 5.4 | 26.02 | 14.26 | B | 1995 |
| Germany | 82.8 | 27.01 | 22.25 | B | 1998 |
| UK <sup>(1)</sup> | 66.4 | 29.01 | 80.92 | B3 | 2005 |
| Finland | 5.5 | 29.01 | 6.18 | B | 2006 |
| France <sup>(1)</sup> | 67.0 | 24.01 | 133.0 | B3 | 2007 |

\* Second wave cases
<sup>(1)</sup> First vaccination at older age

Apart from Wave 2 cases that are likely still developing, the only noticeable outliers in the clear trend: the lower m.p.c., the later happened the cessation of the UIP are Australia; United Kingdom; and France.

The case of Australia needs to be analyzed in more detail; is it a statistical fluctuation, an artefact of reporting mechanism; or some other influencing factor?

The other two outlier cases, UK and France have in common 1) a strong deviation toward much higher level of m.p.c. than the trend; and 2) the administration practice of BCG, namely not at birth but at an older age, as late as school for UK and possibly, France as well. This observation provides the ground for the age effectiveness hypothesis, see sections 4.2 and 5 for further discussion.

#### 7. Specific cases of interest

A number of natural cases for which some data is already available can be useful in the analysis of the hypothesis of the study, such as:

Core Europe: Germany – France – UK

Portugal – Spain

Northern Europe (Denmark, Finland, Norway)

Canada: regional variation

Ukraine: regional variation

Canada (Quebec): age cohort variation

New York – Kyiv

The effective Asian group (Japan, South Korea, Taiwan, Singapore).

Some of these cases are discussed further in the following section.

## 4 Case Analysis

Further studies on statistical significance of noted examples are needed. Some directions can be suggested in which it may be possible to attain conclusive results with the analysis of already available data.

### 4.1 Global Coverage Analysis

A good collection of data on BCG immunization is available in the BCG World Atlas [4]. With the available data, a subset of countries with consistent and widespread immunization policy, for example, on levels A and B as defined above, can be determined and the effects of Covid-19 in spread and severity evaluated. If most of these jurisdictions indeed would show the mild pattern of propagation of Covid-19, it would strengthen the case for the correlation substantially.

A first attempt at such analysis based on preliminary and almost certainly incomplete data that was available at the time of writing was attempted in Table 1. Ideally, a map of such data can be kept and updated continuously, as a ground for more reliable analysis.

### 4.2 Core Europe

The difference between Germany on one hand, and France and UK on the other (Table 1) is quite puzzling and even astounding. All countries share similar culture, demographics, level of economic and social development, traditions and even as can be surmised to a significant degree, genetic background. While differences in public health systems and policy making may exist, we find it difficult to expect that such a large gap can be attributed solely to the policy and administration area. This leaves the possibility of an additional and significant factor at play in this case. Turning to the immunization record for these countries [4]:

Germany: UIP 1961 – 1998 (East Germany probably from 1950-s), administration: at birth. Age cohorts not covered by UIP are 1) 20 or under; and 2) over 60.

UK: UIP 1953 – 2005, administration: school age (12-13). Age cohorts not covered by UIP: 1) 15 and under; 2) over 68. Note that immunization was given not at birth, but a school age.

France: 1950 – 2007, administration: needs verification. Age cohorts not covered by UIP: 1) 13 and under; 2) over 70. Due to a contradiction the immunization age policy needs to be verified further: while [3] indicates school age, [4] states at birth.

Italy: UIP was never offered.

While further analysis is needed for this group one can note that the available Covid-19 impact statistics are consistent with the hypothesis of an additional protection from UIP/BCG <u>with effectiveness further dependent on the age of administration</u>. For example, countries where UIP was offered for an extended period at an older age (UK and possibly France), all in the first wave, are showing m.p.c. rates (65 – 100) right between those of countries with UIP immunization at birth (groups VL and L) and those that never had UIP (m.p.c. > 200).

### 4.3 Regional and Age Cohort Variation Analysis

In certain jurisdictions significant regional variability in administration of BCG vaccination can be noted.

#### Portugal – Spain

An interesting case of a regional variation is Portugal vs. Spain. Geographical neighbors sharing similar traditions and culture, the difference between the impacts of the epidemics is quite dramatic: m.p.c. of 25.8 (Portugal) vs. 253 (Spain) i.e. close to an order of magnitude! Unlike Spain, Portugal has a UIP against tuberculosis with BCG.

On the other hand, the timing of introduction appears to be different in these countries (Portugal: wave 2, Spain: first wave) so a definitive determination of the scenario in Portugal can be made at a future time, approximately TZ + 3.5m.

#### Canada: Regional Variation

In Canada, while there was no consistent effort of universal vaccination on the national level, in some regional jurisdictions it was administered in some age cohorts (e.g. Quebec, 1956-74, [5]) or groups of indigenous population with higher incidence of tuberculosis [6]. Comparing these variations with the epidemiological statistics on Covid-19 in the corresponding age and /or specific groups can shed further light on the case for the correlation. For example, Prairie provinces have significantly higher proportion of the indigenous population where immunization is more widely used than in the other areas of Canada also have significantly lower statistics of Covid-19 at this time according to the results in Table 1, though timing factor can be essential as well due to delayed introduction. Again, a more definitive determination can be made in 2-3 weeks, i.e. ∼ TZ + 3.5m.

#### Ukraine: Regional Variation

In some jurisdictions of the former Communist block such as Ukraine, the scenario can be opposite. While universal BCG has been administered here universally to the current day [4], some regions for example in Western Ukraine for historic or traditional reasons, may have varying degree of the coverage. Anecdotally, the same regions appear to be showing higher effects of the epidemics in both cases and mortality [7], however again these cases need to be carefully validated statistically.

#### Northern Europe

The interesting observation that can be made about this group of four countries: Sweden, Norway, Finland and Denmark, is the large variation in the impacts of Covid-19 with very similar factors including BCG immunization with m.p.c. ranging from 0.22 (Denmark) to 37.8 (Sweden). While situation may still be developing in the countries of the second wave (Denmark, Norway) and statistical fluctuations are possible (see p.4, Section 3.2) such a variation may ask for a more in-depth analysis of the difference that could explain these statistics.

Update: later data revealed that the case of Denmark was a statistical or reporting fluctuation in the developing situation and a general trend between later cessation of UIP and lower Covid-19 mortality is well expressed in this group.

#### Age Cohort Variation: Quebec

An interesting natural experiment opportunity is presented by Quebec universal BCG immunization program administered in 1956-74 [5]. When sufficient Covid-19 statistics are collected it would be possible to compare the impact of Covid-19 on the immunized cohort (aged 45 – 65, at the time of writing) with the adjacent ones (65 – 85) and (25 – 45) by vulnerability to Covid-19 infection, i.e. the infection rate and the severity of outcomes.

#### USA: Regional

The authors are not aware at this time of comprehensive data on regional and local BCG immunization records for the United States but due to wide regional variety interesting cases for the study of the link may exist as well. Of special interest would be any records of local or regional immunization effort, universal or widespread.

### 4.4 New York vs Kyiv

The cities offer almost a classical case for a comparison analysis with a very clear separation by most factors. Large and dense urban centers with high concentration of population, a large network of public transport, bustling social, commercial and entertainment hubs. Both are busy and popular hubs of national and international travel. Since 2017 Ukraine has a visa-free travel agreement with the European Union.

#### Known factors

Demographics: most vulnerable to Covid-19 demographics are strongly represented in both cities, though more detailed study may be needed

Social, tradition, lifestyle: close social links with entertainment and social communication common and widespread

Policies, economic development: NYC is in the high income / development category; Ukraine on the other hand has a lower development rating though Kyiv, the capital is and likely by far, at the highest income and development level in the country.

Quality and efficiency of the healthcare system: most likely more advanced in NYC, more detailed study is needed

Quality of policy making and execution: restrictions related to Covid-19 were implemented at an earlier stage in Kyiv and were by far more extensive. For example, public transport in Kyiv was not available for the general population as of LTZ+1w while subway system was closed on LTZ+1d in Kyiv.

#### Time of introduction

LTZ in New York is: 01.03.2020 (wave 2)

LTZ in Kyiv is: 16.03.2020 (wave 2)

While both cities are in the wave 2 of the pandemics, the difference in two weeks strongly matters for Covid-19 statistics especially during the period of onset. The situation should be revisited at the similar phase of development in Kyiv i.e. ∼ LTZ + 1m ∼ TZ + 3.5m.

#### Covid-19 Impacts

NYC total mortality / m.p.c. as of LTZ + 1m: 2254 / 262.1

Kyiv total mortality / m.p.c. as of LTZ + 0.5m: 4 / 1.05.

NYC / Kyiv, total mortality: 563.5

NYC / Kyiv, m.p.c.: 249.6

If the current trend is maintained within the next two weeks i.e. up to the LTZ + 1m (Kyiv) = TZ + 3.5m it could be one of the strongest cases for the factor F hypothesis.

### 4.5 The Effective Asian Group

All countries in this group (Japan, South Korea, Taiwan, Singapore) are:

- The first wave countries, with the longest and likely, most reliable statistics
- All countries are in the “high income” development group
- All countries have noticeably less severe Covid-19 impacts than most of the affected jurisdictions, with m.p.c. in the range of 1% or even significantly lower (Taiwan: 0.2).
- All countries in the group have strong Covid-19 prevention policies developed from the experience of the previous incidents (SARS-2002 [8], MERS [9])
- All countries have a current UIP/BCG.

The example of this group, certainly not an extraordinary outlier, clearly show that the combination of an effective prevention strategy and policies combined with possibly, the protective effect of a universal immunization can go a long way to reducing the negative impacts of the Covid-19 epidemics on the population.

## 5 Possible Mechanism

Based only on a remote and unverified at this time connections, one may come with a conjecture that the effectiveness of the protection caused by the immunization factor, if confirmed, over the lifetime is controlled by a formation of some immune protection mechanism in the early development. Such a supposition is based on the following connections:

- The analysis in Section 4.2 of otherwise equal jurisdictions appears to suggest that the early age immunization may offer more effective protection against Covid-19 over the lifetime.
- BCG vaccine has uses for immunotherapy [10] and is known to provide some level of support for immune system in a number of conditions.
- Some studies previously indicated higher protective effect of the earliest delivery of the vaccine against tuberculosis [11]
- A gender link in the immune protection against Covid-19 has been suggested previously [12]. Such a gender-specific mechanism, if confirmed, would likely require a development from an early age, that though not necessarily the same expression, may be also true for the purported mechanism of BCG-triggered protection.

On the basis of these associations a conjecture can be proposed that an early exposure to a vaccine may provide the immune system with a cue to develop or emphasize certain functions and capacities that would be more effective in protecting the organism against an attack of the virus over the lifetime. At this time this only a hypothesis that will need further research to verify or disprove.

## 6 Conclusion

We believe that the data compiled in this work as the early observations obtained with it can be useful to other researchers in the field looking for effective approaches to control the pandemics. In the authors view these early results offer more convincing arguments in support of the hypothesis of BCG immunization protection effect against Covid-19 providing a strong rationale for further studies that would result in a confident judgement.

As well, in our view, the hypothesis of the early age effectiveness for the protection has certain arguments in its support and deserves further attention and study that perhaps may lead to a direction toward developing effective immune means to protect against the infection and control it.

## Data Availability

All data referred to in the paper was available online at the time of submission

https://www.google.com/covid19-map/

http://www.bcgatlas.org/

## Notes

### Competing Interest Statement

The authors have declared no competing interest.

### Funding Statement

This work was not supported by any funding.

